# Heterogeneity in susceptibility among humans to common respiratory viral infections

**DOI:** 10.64898/2026.05.29.26353692

**Authors:** Kaho Shinozaki, Fuminari Miura

## Abstract

**Background:** Human challenge trials provide a unique opportunity to quantify pathogen infectivity in terms of the probability of infection given an inoculated dose. However, between-pathogen comparisons are often distorted by individual heterogeneity in host susceptibility and by differences in background immunity across trial populations. We examined how dose–dependent infection risks differ across common respiratory viruses when such heterogeneity is explicitly incorporated.

**Methods:** We conducted a systematic review of human challenge trials for four respiratory viruses: respiratory syncytial virus (RSV), influenza virus, rhinovirus, and adenovirus. Using the extracted data, we fitted dose–response models under different distributional assumptions, allowing both continuous susceptibility variation and discrete immune fractions. We compared alternative heterogeneity models and evaluated pathogen-specific dose–response patterns using original and scaled dose metrics.

**Results:** All four viruses showed substantial heterogeneity in host susceptibility, and models assuming homogeneous susceptibility were unsupported. RSV and influenza were best described by models with a distinct immune or effectively non-susceptible subgroup, and the estimated immune proportions were approximately 40% and 25%, respectively. In contrast, rhinovirus and adenovirus were better explained by continuously distributed susceptibility, with little evidence of a fully immune subgroup. On a scaled dose axis, rhinovirus and adenovirus showed steeper increases in infection risk with dose than RSV and influenza.

**Conclusions:** The structure of susceptibility heterogeneity differs across common respiratory viruses, which in turn shapes dose–dependent infection risks. Incorporating this heterogeneity is essential for valid cross-pathogen comparison and for interpreting human challenge data in epidemiologic and public health contexts.

**Summary:** We systematically reviewed human challenge trials and analyzed the extracted data using dose–response models for four respiratory viruses. The analyses revealed between-pathogen differences in dose–dependent infection risks and highlighted substantial heterogeneity in host susceptibility, even under controlled exposure conditions.

## Background

Respiratory pathogens share common transmission routes, such as droplets and aerosols typically in close-contact settings, yet their transmission patterns differ substantially. These differences are often attributed to population immunity, pathogen-specific immune duration, and seasonal, social or environmental drivers [1,2]. Another key factor to explain this variation is per-exposure infectiousness, that is, the probability that a single exposure results in infection. For respiratory viruses such as respiratory syncytial virus (RSV), influenza virus, rhinovirus, and adenovirus, such differences may contribute to variation in observed transmission dynamics. However, in natural transmission settings, this quantity is difficult to estimate because the effective exposure dose is unmeasured and unobservable.

Human challenge studies have provided a unique experimental framework to overcome this limitation [3–5]. In these controlled experiments, both the dose and route of viral exposure are standardized, enabling direct estimation of pathogen-specific dose–response relationships. By quantifying infection probability across increasing exposure levels, challenge studies offer a basis for comparing intrinsic infectiousness across pathogens. However, even under controlled exposure conditions, infection outcomes vary substantially among individuals, reflecting inter-individual heterogeneity in susceptibility [6,7].

Such heterogeneity arises from multiple sources, including prior infection, partial or cross-protective immunity, age, genetic factors, and baseline health status. Some individuals may be effectively resistant to infection, whereas others remain highly susceptible. Failure to account for this heterogeneity can bias estimates of infectiousness and obscure meaningful differences between pathogens [8–10]. To address this challenge, our previous work [11], together with that of others [12,13], introduced dose–response models that explicitly incorporate individual-level heterogeneity in susceptibility, including the possibility that a fraction of individuals may be completely resistant to infection. These models are particularly useful for analyzing human challenge trial data, where participants’ pre-existing immunity is often unknown or incompletely characterized. However, existing dose–response studies have focused on analyses of single pathogens [14]. Characterizing between-pathogen differences in susceptibility structure and infectiousness requires a unified comparative approach that can account for differential background immunity across trial populations.

In this study, we sought to determine how per-exposure infectiousness differs across common respiratory viruses when host susceptibility heterogeneity is explicitly accounted for. To this end, we conducted a systematic review of human challenge studies for four key respiratory viruses (RSV, influenza virus, rhinovirus, and adenovirus) and analyzed the extracted data within a unified dose–response framework that allows for both continuous susceptibility variation and discrete immune fractions. This design allows us to compare pathogens while correcting for differences in background immunity, to characterize infection risks on a unified dose scale, and to assess how inferred infectiousness and susceptibility depend on assumptions about pre-existing immunity among trial participants.

## Methods

### Systematic review of human challenge studies

We performed a literature review to compile publicly available data from human challenge studies involving RSV, rhinovirus, adenovirus, and influenza virus. Study identification followed the Preferred Reporting Items for Systematic Reviews and Meta-Analyses (PRISMA) 2020 guidelines (https://www.prisma-statement.org/prisma-2020-flow-diagram). Literature searches were conducted in PubMed and Ovid MEDLINE for studies published up to September 2025, with no restrictions on publication years. For each pathogen, we searched for published studies with defined and documented doses of each virus and both the number of participants exposed and infected available. Each search used common variations of the virus (e.g., “RSV”, “respiratory syncytial virus”, “RV”, “HRV”, “rhinovirus”, “human rhinovirus”, “AdV”, “hAdV”, “adenovirus” “human adenovirus”, “influenza”, “flu”, and “influenza virus”), combined with terms related to human challenge experiments. The full search strategies for PubMed and Ovid MEDLINE are provided below:

**PubMed**

((“human challenge”[Title] OR “controlled human infection”[Title] OR “challenge study”[Title] OR (experiment*[Title] AND (infect*[Title] OR inoculat*[Title]) AND (human[Title] OR adult*[Title] OR volunteer*[Title])) OR (challenge*[Title] AND (human[Title] OR adult*[Title] OR volunteer*[Title])))) AND ((pathogen[Title]))

**Ovid MEDLINE**

1. (“human challenge” or “controlled human infection” or “challenge study” or (experiment* and (infect* or inoculat*) and (human or adult* or volunteer*)) or (challenge* and (human or adult* or volunteer*))).ti
2. (pathogen).ti
3. 1 AND 2

Note: Here, **pathogen** denotes the virus-specific search string, consisting of all relevant naming variants for each pathogen (e.g., “RSV”, “respiratory syncytial virus”), combined using OR.

All titles and abstracts were manually screened by a single reviewer to identify potentially relevant studies, and full-text reviews were conducted when necessary. Studies were excluded if they were not written in English, were not conducted against human or healthy adult volunteers, did not report explicit inoculation doses, or did not provide sufficient information on infection outcomes. Studies reporting challenge doses only as ranges were also excluded to ensure comparability of dose–response analyses. In addition, we reviewed reference lists of included articles and relevant studies from the authors’ previous publications to identify further eligible publications. A summary of included studies is provided in the e-appendix (GitHub).

Rather than extracting summary statistics, we collected raw reported data from each eligible study. Specifically, the collected quantitative information included: publication year; cohort size; the age range of participants who were challenged; the number of participants who were challenged; the number of challenged participants who became infected; the number of challenged participants who became symptomatic (if available); and the dose of inoculated virus. When virus inoculum doses were reported using different units, all values were standardized to plaque-forming units (PFU) to ensure comparability across studies. For studies reporting viral titers as the 50% tissue culture infectious dose (TCID₅₀), values were converted to PFU using an estimation factor of 0.7 [15]. We also extracted the following qualitative information: title and first author of the study; pathogen (and strain) inoculated; definitions used for infection; previous history of infection or vaccination (if available).

### Dose–response models for human challenge data

To quantify per-exposure infection risk in human challenge studies, we modeled infection probability as a function of the administered challenge dose. The reported infectious dose 𝑑 represents the expected value of a Poisson process describing the number of infectious particles effectively received by a participant, with mean 𝑑. Under the assumption of homogeneous host susceptibility (i.e., all hosts have equal susceptibility) and independent action of infectious particles, the probability of infection after exposure to dose 𝑑 is given by 𝑃(𝑑) = 1 − 𝑒𝑥𝑝(−𝑑). To account for individual variation in susceptibility, we introduced a susceptibility factor 𝑠 for each participant, drawn from a distribution 𝑓(𝑠). A participant with susceptibility 𝑠 has an infection probability 𝑠-fold higher than an individual with 𝑠 = 1 per infectious particle. Under this formulation, the resulting infection probability for a given challenge dose 𝑑 is:

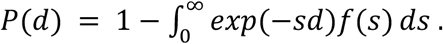

By fitting this model to the observed infection proportions from the human challenge studies, we inferred the shape of the susceptibility distribution 𝑓(𝑠).

We evaluated four candidate models for the susceptibility distribution 𝑓(𝑠): a **Dirac delta distribution**, corresponding to uniform, homogeneous susceptibility among individuals; a **gamma distribution**, representing continuously distributed susceptibility; a **two-level distribution,** assuming two distinct groups with different susceptibility levels 𝑎_1_ and 𝑎_2_, present at proportions 𝑝_1_ and 1 − 𝑝_1_, respectively; and a **gamma distribution with an additional point-mass (gamPM)**, assuming a fraction of the population 𝑝_1_ with homogeneous susceptibility at the level 𝑎_1_, while the remainder (1 – 𝑝_1_) follow a gamma-distributed susceptibility. A variant allowing for a point-mass at zero susceptibility (i.e., 𝑎_1_ is fixed at zero), representing a completely immune subgroup (**gamPM0**), was also considered.

### Parameter estimation, uncertainty quantification, and scaled comparisons of inferred dose–response curves

For each pathogen and candidate model, we derived the binomial likelihood functions based on the observed number of infected and uninfected participants at each challenge dose. Model parameters were estimated by the maximum likelihood estimation, fitting each dose–response model to each pathogen-specific dataset. Model fit was assessed using the Akaike Information Criterion (AIC), and models were compared based on AIC to identify the susceptibility distribution that best explained the observed infection outcomes for each virus.

Uncertainty in parameter estimates was assessed using a parametric bootstrap procedure with 1000 iterations. For each fitted model, infection outcomes were resampled from a binomial distribution with trial size equal to the number of challenged participants and success probability given by the fitted dose–response function. The model was re-fitted to each bootstrap replicate, and 95% confidence intervals were derived from the 2.5th and 97.5th percentiles of the resulting parameter distributions.

### Comparative characterization of susceptibility distributions

To compare the distributional properties of host susceptibility across pathogens, we examined summary statistics derived from fitted susceptibility distributions. For each pathogen and for models allowing heterogeneous susceptibility (two-level, gamma, gamma plus point-mass, and gamma plus point-mass at zero), we computed key distributional measures, including the mean, variance, coefficient of variation (CV), and, where applicable, the estimated proportion of immune individuals. These quantities were jointly visualized using scatter plots to facilitate descriptive comparison of characteristics of the susceptibility distribution across pathogens and model structures.

Since the challenge dose units are not always comparable across pathogens (e.g., 1 TCID50 for influenza and RSV does not necessarily represent equivalent infectious exposure), we examined scaled dose–response curves in which mean host susceptibility was normalized to one. This normalization allows comparison of curve shapes without relying on absolute equivalence of dose units across studies or pathogens. Under this condition, differences in dose–response relationships can be interpreted in terms of how rapidly infection probability changes with increasing dose, providing a standardized representation of dose sensitivity across pathogens.

Because immune or low-susceptibility subgroups can influence the interpretation of infection probabilities, we additionally examined dose–response curves adjusted for the estimated proportion of immune individuals. For each pathogen, infection probabilities estimated from the fitted models were recalculated by correcting for the immune fraction (i.e., by setting the immune proportion parameter to zero). This yields dose–response relationships corresponding to a fully susceptible population. These adjusted curves were compared with the original (unadjusted) curves to evaluate the impact of ignoring immune subgroups on inferred dose–response relationships.

## Results

### Study selection

A total number of 121 studies were included: 22 for RSV, 17 for rhinovirus, 4 for adenovirus, and 78 influenza virus, respectively. **Figure 1** shows an aggregated PRISMA flow-chart of study selection, while the flowcharts for each pathogen and the table of the reviewed articles (for each pathogen) are provided in **Figures S1-S4** and the **e-Appendix (GitHub)**, respectively. Of the 121 studies, 66 were retrieved from two databases, and 55 were identified from citations.

**Figure 1.**
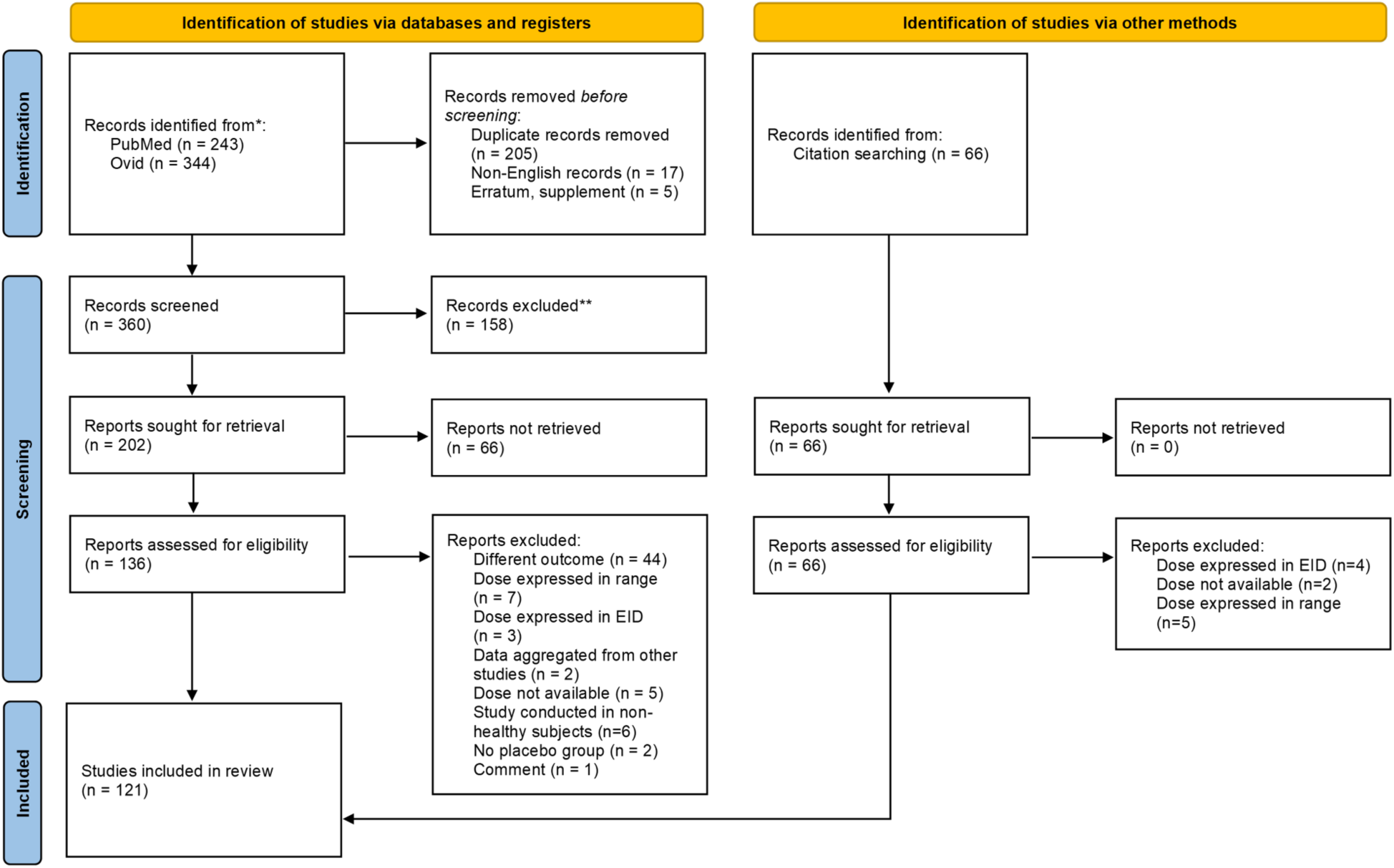
Aggregated PRISMA flow diagram of study selection. Flow diagram summarizing identification, screening, eligibility assessment, and inclusion of human challenge studies for respiratory pathogens in this systematic review. Records were identified through database searches (PubMed and Ovid MEDLINE) and citation screening. After duplicate removal and screening, full-text articles were assessed for eligibility based on predefined criteria, including availability of challenge dose and infection outcome data.

### Inferred dose–response models

The best-fitting models differed across pathogens. The gamma plus point-mass at zero model provided the best fit for RSV and influenza virus, whereas the gamma model was selected as the best model for rhinovirus and adenovirus (**Figure 2**). These findings imply that there may exist a non-susceptible (immune) subgroup among the study participants for RSV and influenza virus, but not for rhinovirus and adenovirus. For all pathogens, models assuming homogenous levels of susceptibility were not supported by the data. Model comparison results based on AIC for each model by pathogen are summarized in **Table S1**, and fitted dose–response curves for each model by pathogen are shown in **Figures S5-S8**.

**Figure 2.**
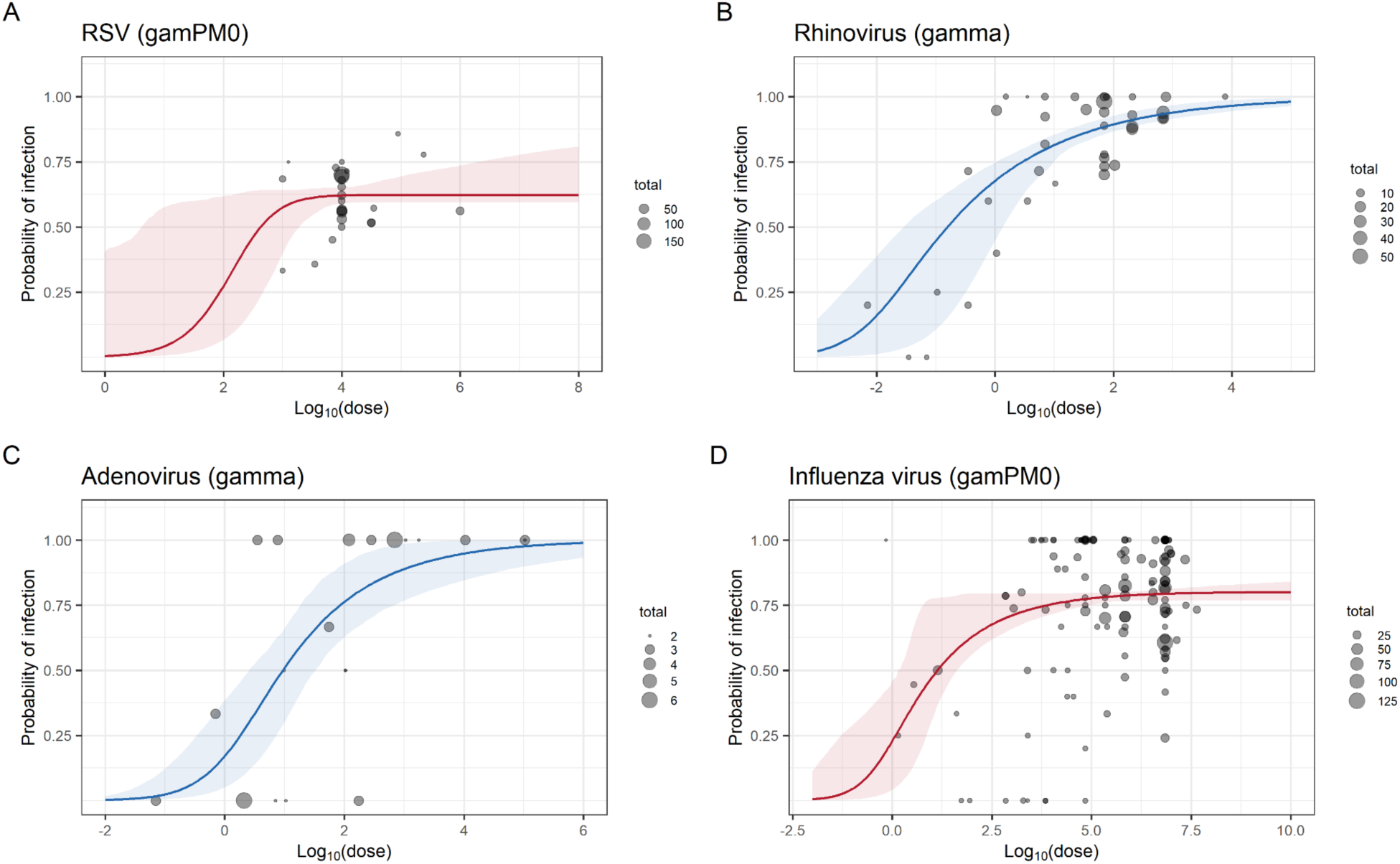
Best-fitting dose response curves for each pathogen based on observed human challenge data. Grey bubbles represent observed data, with circle size proportional to the number of participants in each trial at each dose level. Colored curves show fitted dose–response models (blue: gamma model; red: gamma plus point-mass at zero model). Panels display the best-fitting model for each pathogen: (A) RSV, gamma plus point-mass at zero model; (B) rhinovirus, gamma model; (C) adenovirus, gamma model; (D) influenza virus, gamma plus point-mass at zero model.

### Comparison of susceptibility distributions

Summary statistics for each pathogen-model combination are shown in **Figure 3** to compare the characteristics of susceptibility distributions across pathogens and modeling assumptions. Figure 3(A) shows that RSV has lower mean and variance than other pathogens in all models, consistent with the higher inoculation doses required to establish infection with RSV. In **Figure 3(B)**, the two-level model shows consistent estimates for all pathogens, with the smallest mean and CV among heterogeneous susceptibility models (i.e., except for the delta model). The gamma models for influenza produced higher mean, variance, and CV values compared to other models (**Figure 3(A)** and **3(B)**), suggesting that gamma models might overestimate the mean and variance owing to an assumption of no immune proportion. The estimated immune proportion 𝑝_1_ is shown in **Figure 3(C)**. For RSV, approximately 30-40% of individuals were inferred to be non-susceptible, whereas no immune fraction was detected for adenovirus across all models.

**Figure 3.**
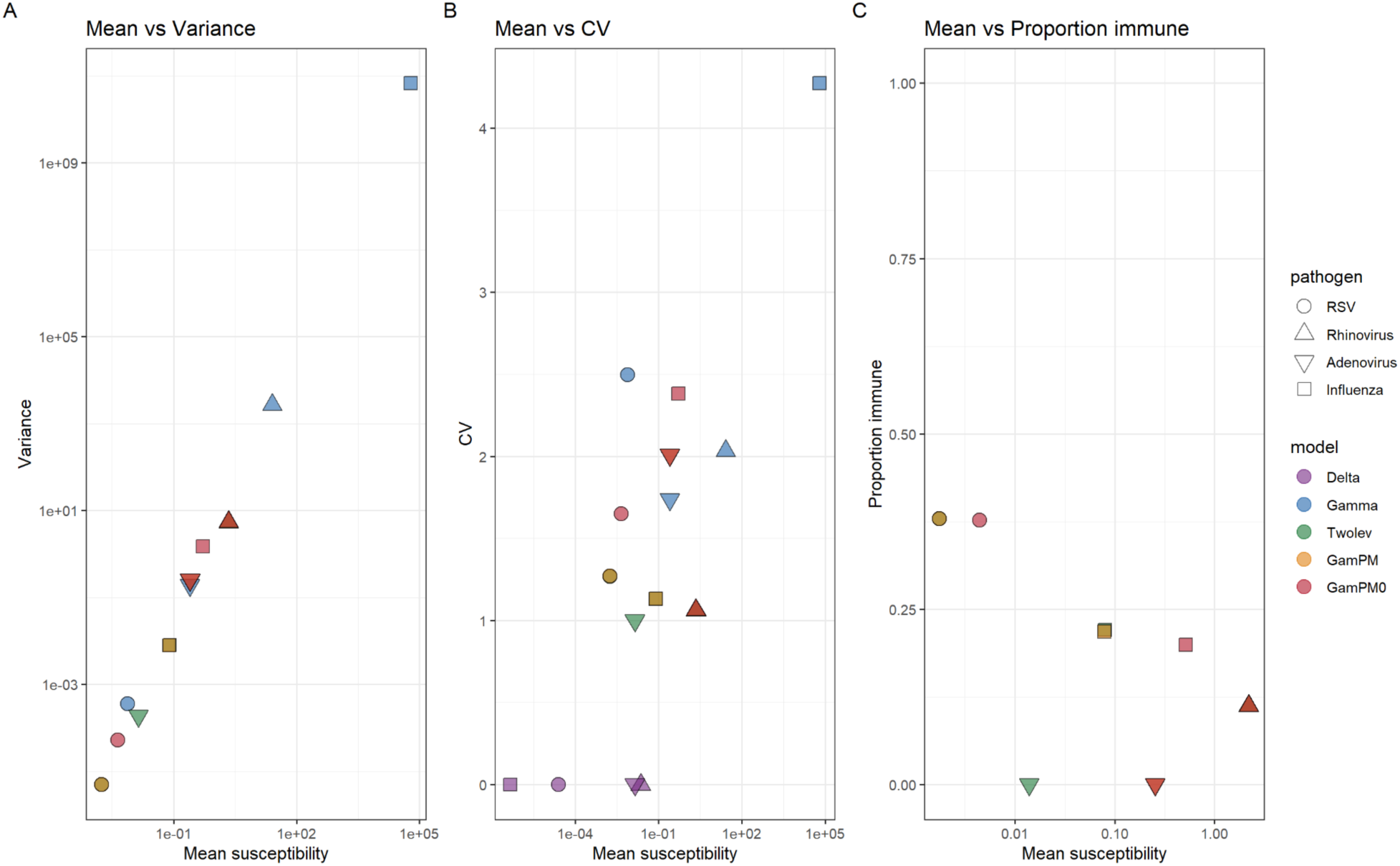
Comparison of inferred susceptibility distributions across pathogens and models. Scatter plots show the relationship between the estimated mean susceptibility (x-axis) and (A) variance, (B) coefficient of variation (CV), and (C) the proportion of immune individuals. Each point represents the inferred susceptibility distribution for a given pathogen–model combination.

### Scaled dose–response curves

Scaled dose–response models were additionally used to compare the sensitivity of infection probability to dose (i.e., the rate at which infection probability increases by dose) across pathogens (**Figure S9**). In the scaled gamma model (**Figure S9(A)**), the curves rise most steeply and reach 100% infection at the lowest doses in the order of adenovirus, rhinovirus, RSV, and influenza virus. In the scaled gamma plus point-mass at zero model (**Figure S9(B)**), the relative order of curves remains similar at higher doses, with adenovirus and rhinovirus displaying left-shifted curves and comparatively steeper at lower doses. These results indicate that infection probability increases more rapidly with dose for these two viruses compared with RSV and influenza virus.

### Dose–response curves adjusted for the estimated immune proportion

Dose–response curves derived from the gamma plus point-mass at zero model (gamPM0) are shown in **Figure 4**. For adenovirus, the estimated infection probability increased monotonically with dose and approached the value of one without any adjustment for an immune fraction, suggesting that continuous heterogeneity in susceptibility alone (gamma distribution) is sufficient to explain the observed infection outcomes. In contrast, for RSV, rhinovirus, and influenza virus, the estimated dose–response curves exhibited a plateau below an infection probability of one unless the immune proportion parameter (𝑝_1_) was constrained to 0. The estimated immune fractions were approximately 40% for RSV, 10% for rhinovirus, and 25% for influenza virus, consistent with the estimates shown in **Figure 3C**.

**Figure 4.**
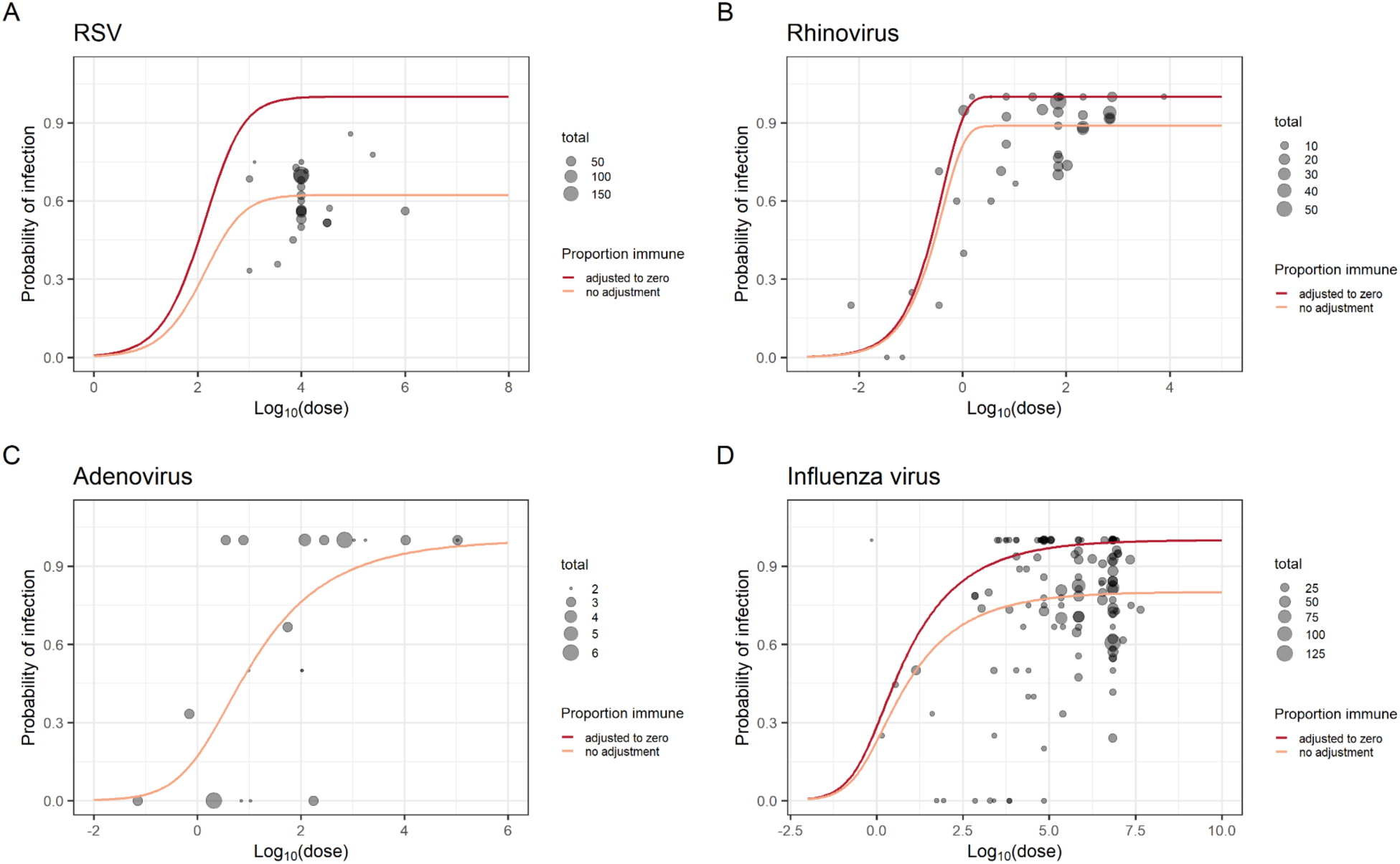
Fitted dose–response curves for each pathogen under the gamma plus point-mass at zero model. Grey bubbles represent the observed data, and their sizes represent the number of participants in each trial at each dose. Orange lines indicate fitted dose–response curves without any adjustment, and red lines indicate curves corrected for the estimated proportion of immune individuals. Panels correspond to (A) RSV, (B) rhinovirus, (C) adenovirus, and (D) influenza virus.

The discrepancy between the adjusted and unadjusted dose–response curves was pronounced particularly for RSV and influenza virus. Whereas the unadjusted curves were bounded at infection probabilities substantially lower than one, the adjusted curves continued to rise toward the infection probability of one (due to the removal of immune individuals). This divergence was most evident at higher dose ranges, where additional increases in dose produced little change in the unadjusted curves but resulted in much higher estimated infection probabilities after adjustment.

## Discussion

In this study, we compared per-exposure infectiousness across four common respiratory viruses using human challenge data and dose–response models that explicitly account for heterogeneity in host susceptibility. Our analyses demonstrate that both the shape of dose–response curves and the underlying variation in susceptibility differ substantially across pathogens. Models assuming homogeneous susceptibility were unsupported for all viruses examined, indicating that inter-individual variation is a defining factor of infection risk even under controlled exposure conditions.

The comparative results highlight pathogen-specific patterns of susceptibility. For RSV and influenza virus, the best-fitting models included a distinct immune or non-susceptible subpopulation, with estimated immune fractions of approximately 40% for RSV and 25% for influenza, respectively. The corresponding dose–response curves plateaued below an infection probability of one. This implies that increasing the inoculum dose does not necessarily lead to a proportional increase in infection risk, as a subset of participants remains effectively protected against infection. These findings are biologically plausible given the high prevalence of prior infection and the role of partial or cross-reactive immune protection [16–18]. In contrast, rhinovirus and adenovirus challenge data were best explained by continuously distributed susceptibility (i.e., gamma models), with little evidence for a completely immune subgroup. This pattern is consistent with the fact that immunity is largely serotype-specific for both rhinovirus and adenovirus, where prior infection with one serotype does not typically confer protection against others [19]. Such limited cross-protection may make a near-complete immune subgroup less likely to appear in challenge study populations.

Direct comparison of dose–response relationships across pathogens is challenging because challenge dose units do not necessarily represent equivalent infectious exposures across viruses. To address this limitation, we employed two complementary approaches to interpret and compare dose–response relationships. The first approach adjusts dose–response curves for the presence of an immune or non-susceptible fraction, allowing infection risk among susceptible individuals to be interpreted more accurately by avoiding dilution of the estimated infection probabilities, particularly at higher doses (**Figure 4**). The second approach uses scaled dose–response curves in which mean host susceptibility is normalized to one (**Figure S9**), enabling comparison of curve shapes across pathogens independent of the absolute dose unit. Together, these approaches provide a framework for cross-pathogen comparison even when trial populations differ in baseline immunity and susceptibility structure. On the scaled-dose axis, infection risk increased more steeply for rhinovirus and adenovirus than for RSV and influenza. This in turn suggests that, for a given proportional reduction in exposure, infection risk would decline more markedly for adenovirus and rhinovirus than for RSV and influenza.

This study has several implications for the design and interpretation of HCTs. First, reliance on a single inoculation dose limits the ability to characterize the infection risk and obscures heterogeneity in susceptibility. Incorporating two or more challenge doses and reporting explicit doses would greatly enhance the identifiability of the variation in susceptibility and provide an empirical basis for more accurately comparing dose-dependent infection risks. Second, the existence of immune or low-susceptibility subgroups should be anticipated even in pre-screened cohorts [20], as antibody-based screening does not fully capture mucosal immunity, cellular responses, or other protective mechanisms. Ignoring such heterogeneity may lead to underestimation of infection risk, particularly at higher doses where attack rates plateau. This consideration is particularly relevant for vaccine efficacy trials using HCT endpoints: if a substantial non-susceptible subgroup is present, as suggested for RSV and influenza, standard attack-rate-based efficacy estimates may partly reflect the baseline susceptibility structure of the trial population, not solely the vaccine’s protective effect. Third, our systematic review highlights a clear imbalance in the availability of HCT data across respiratory pathogens. While a large number of challenge studies have been conducted for the influenza virus for the evaluation of vaccines and antiviral compounds [14], only four adenovirus HCTs were eligible for inclusion in the present analysis. This disparity limits inference on dose–response relationships and susceptibility heterogeneity for this pathogen. Expanding HCTs for underrepresented pathogens and exploring a wider range of inoculum doses would greatly help us quantify per-exposure infectiousness and understand how host susceptibility shapes infection dynamics across respiratory viruses.

Several limitations should be acknowledged. The availability of human challenge data varied substantially across pathogens, with particularly limited data for adenovirus compared with the extensive literature on influenza virus. In addition, most datasets for adenovirus contained dose levels clustered near zero or near-complete infection, limiting the resolution of intermediate dose–response regions. Furthermore, study populations were pooled across different time periods and epidemiologic contexts; more stratified analyses accounting for background prevalence or prior circulation intensity may provide further insight into how population immunity influences the shape of estimated susceptibility distributions. Pooling data across strains also precluded assessment of strain-specific differences in infectivity or immunity. Finally, as this study focused on challenge trials with explicitly defined inoculation doses, studies without precise dose information were excluded, which may have introduced some selection bias. Nevertheless, given the consistency of findings across multiple models and pathogens, we believe that these limitations are unlikely to alter the main qualitative conclusions.

In conclusion, this study demonstrates that heterogeneity in host susceptibility is a defining feature of dose–response relationships in human challenge trials and that its distributional variation substantially differs across respiratory viruses. Even under controlled exposure conditions, per-exposure infectiousness cannot be interpreted independently of host susceptibility distributions. Accounting for this heterogeneity is therefore essential for valid comparison across pathogens and for translating challenge study results into epidemiologic insights, infection control planning, and public health risk assessment.

## Author contributions

K.S and F.M. conceptualized the study and developed the methodology. K.S. developed the systematic review protocol, conducted the literature search, curated data, performed the formal analysis, and wrote the first draft of the manuscript. F.M. supervised the study, validated the result, and acquired funding. All authors reviewed and edited the manuscript and approved the final version.

## Acknowledgements

We thank the authors of the original human challenge studies included in this review for making their data available through published reports.

## Data availability

The analysis code and analyzed data underlying this article are available in the GitHub repository at: https://github.com/kaho12may-ux/RespDR_2025.

## List of supplementary materials

**Table S1.** AICs for each distribution by pathogen

**Figure S1.** PRISMA flow diagram of study selection for RSV

**Figure S2**. PRISMA flow diagram of study selection for rhinovirus

**Figure S3.** PRISMA flow diagram of study selection for adenovirus

**Figure S4.** PRISMA flow diagram of study selection for influenza virus

**Figure S5.** Fitted dose response curves for RSV based on observed human challenge data

**Figure S6.** Fitted dose response curves for rhinovirus based on observed human challenge data

**Figure S7.** Fitted dose response curves for adenovirus based on observed human challenge data

**Figure S8.** Fitted dose response curves for influenza virus based on observed human challenge data

**Figure S9.** Fitted mean-scaled dose response curves for each pathogen based on observed human challenge data

## Supplementary tables

**Table S1.** AICs for each distribution by pathogen.

|  | RSV | Rhinovirus | Adenovirus | Influenza virus |
| --- | --- | --- | --- | --- |
| Delta | 2549.30 | 880.15 | 76.37 | 7281.51 |
| Gamma | 1219.48 | 357.88 | 58.07 | 2377.80 |
| Two level | 1206.92 | 360.10 | 80.37 | 2386.85 |
| gamPM (Gamma + point-mass) | 1208.92 | 362.10 | 62.07 | 2388.93 |
| gamPM0 (Gamma + point-mass at zero distribution) | 1206.82 | 360.10 | 60.07 | 2368.84 |

## Supplementary Figure legends

**Figure S1.**
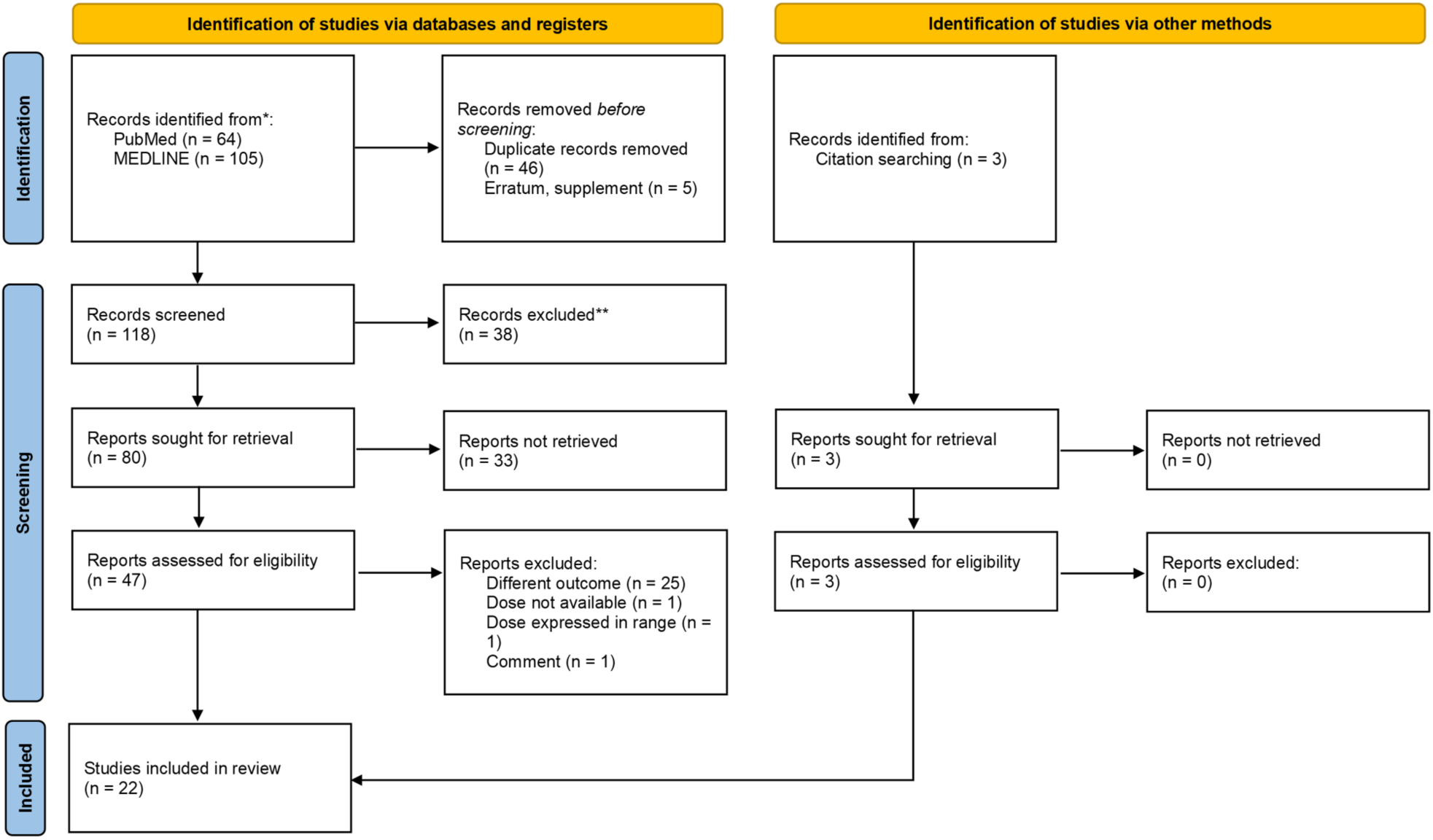
PRISMA flow diagram of study selection for RSV. Flow diagram summarizing identification, screening, eligibility assessment, and inclusion of human challenge studies for RSV in this systematic review. Records were identified through database searches (PubMed and Ovid MEDLINE) and citation screening. After duplicate removal and screening, full-text articles were assessed for eligibility based on predefined criteria, including availability of challenge dose and infection outcome data.

**Figure S2.**
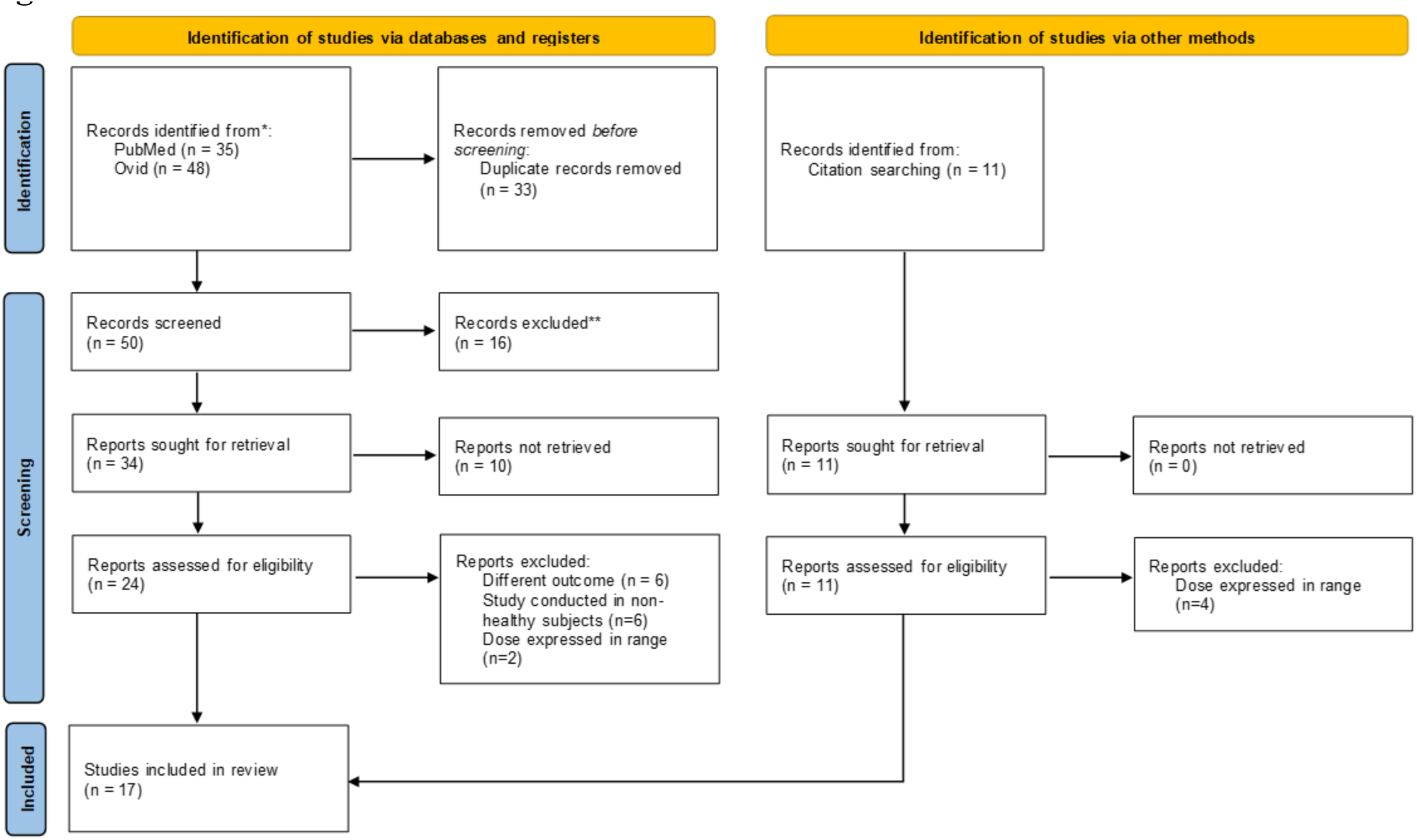
PRISMA flow diagram of study selection for rhinovirus. Flow diagram summarizing identification, screening, eligibility assessment, and inclusion of human challenge studies for rhinovirus in this systematic review. Records were identified through database searches (PubMed and Ovid MEDLINE) and citation screening. After duplicate removal and screening, full-text articles were assessed for eligibility based on predefined criteria, including availability of challenge dose and infection outcome data.

**Figure S3.**
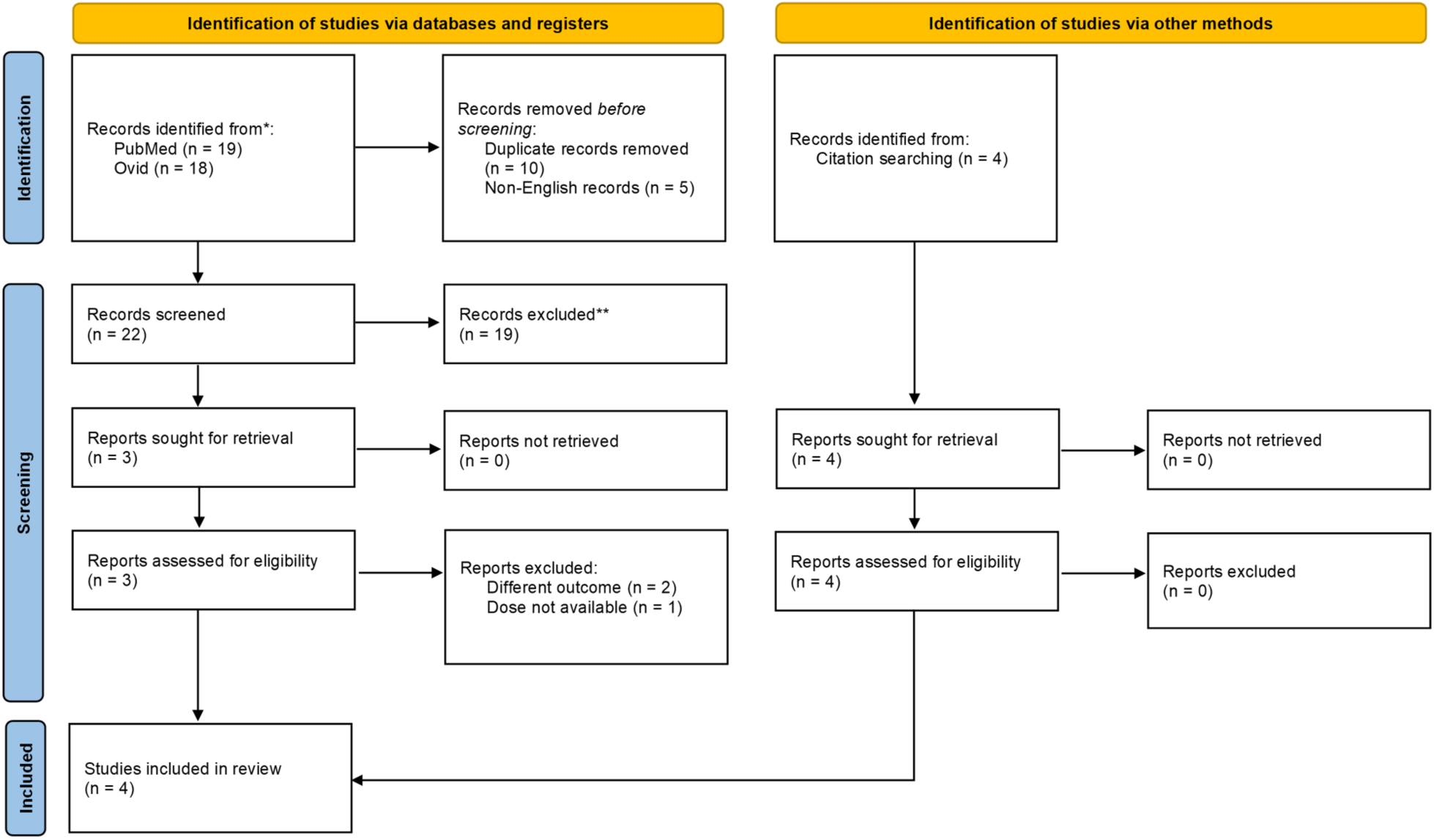
PRISMA flow diagram of study selection for adenovirus. Flow diagram summarizing identification, screening, eligibility assessment, and inclusion of human challenge studies for adenovirus in this systematic review. Records were identified through database searches (PubMed and Ovid MEDLINE) and citation screening. After duplicate removal and screening, full-text articles were assessed for eligibility based on predefined criteria, including availability of challenge dose and infection outcome data.

**Figure S4.**
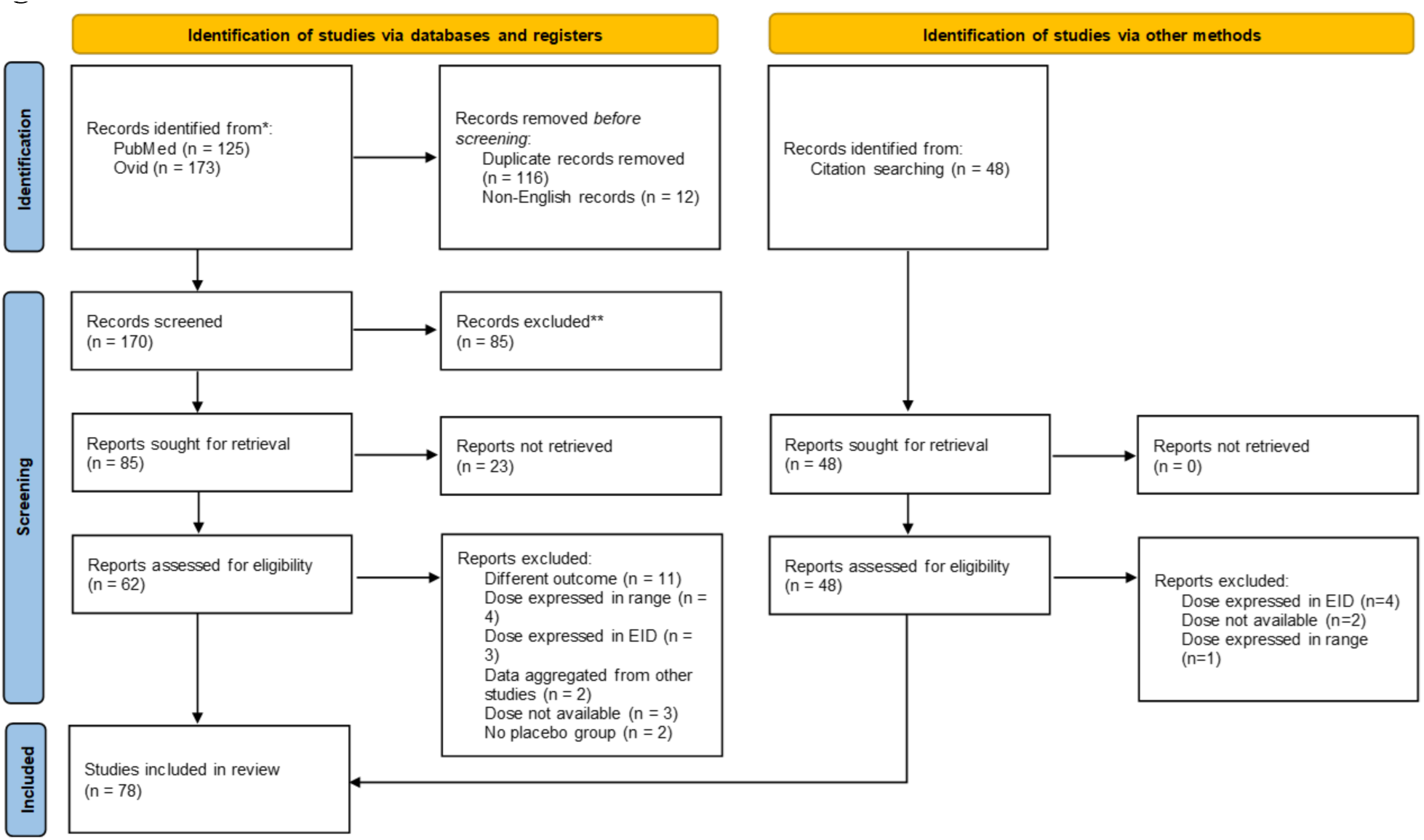
PRISMA flow diagram of study selection for influenza virus. Flow diagram summarizing identification, screening, eligibility assessment, and inclusion of human challenge studies for influenza virus in this systematic review. Records were identified through database searches (PubMed and Ovid MEDLINE) and citation screening. After duplicate removal and screening, full-text articles were assessed for eligibility based on predefined criteria, including availability of challenge dose and infection outcome data.

**Figure S5.**
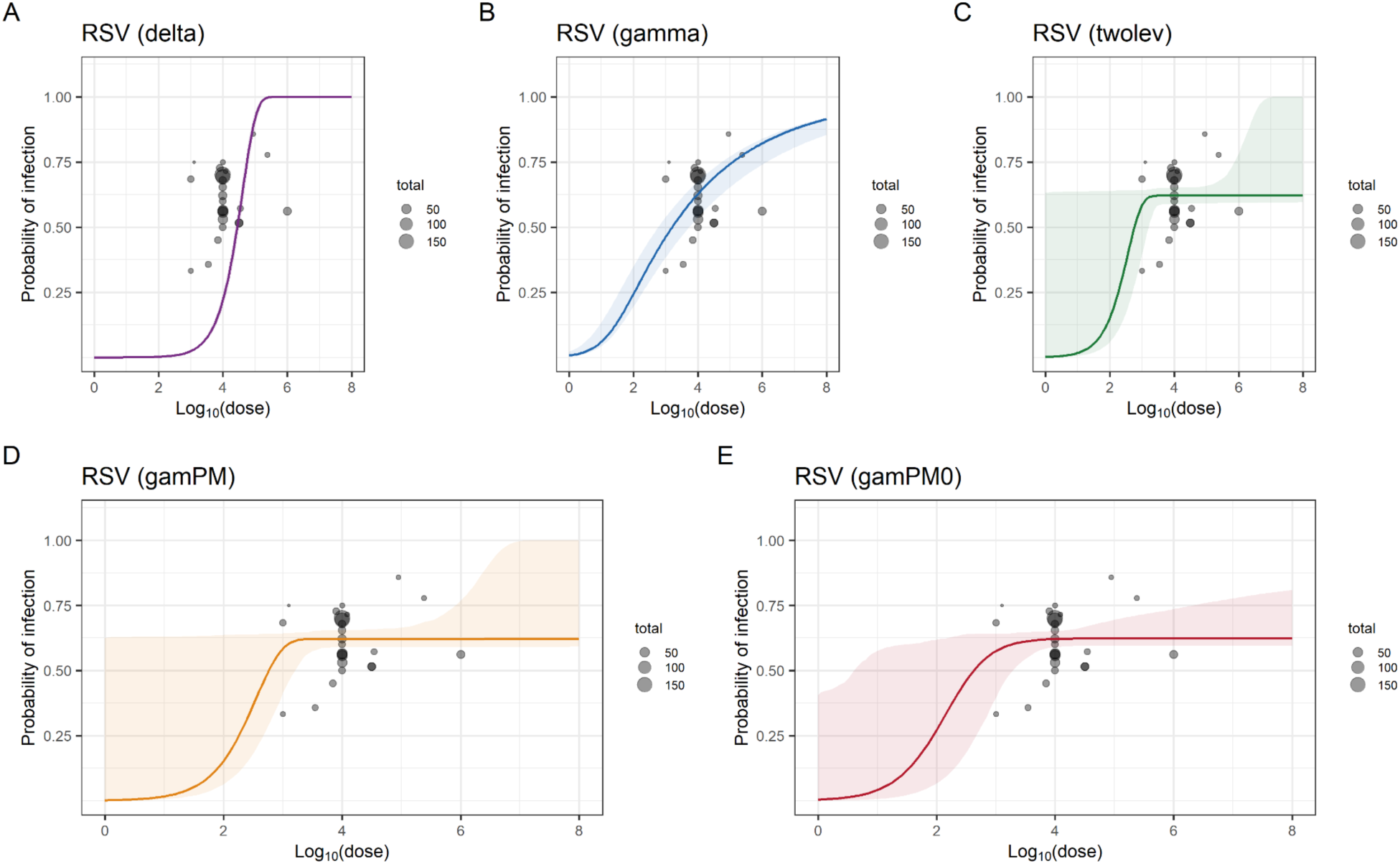
Fitted dose response curves for RSV based on observed human challenge data. Grey bubbles represent observed data, with circle size proportional to the number of participants in each trial at each dose level. Colored curves show fitted dose–response models. Panels display each model for RSV: (A) dirac delta model; (B) gamma model; (C) two-level model; (D) gamma plus point-mass model; (E) gamma plus point-mass at zero model.

**Figure S6.**
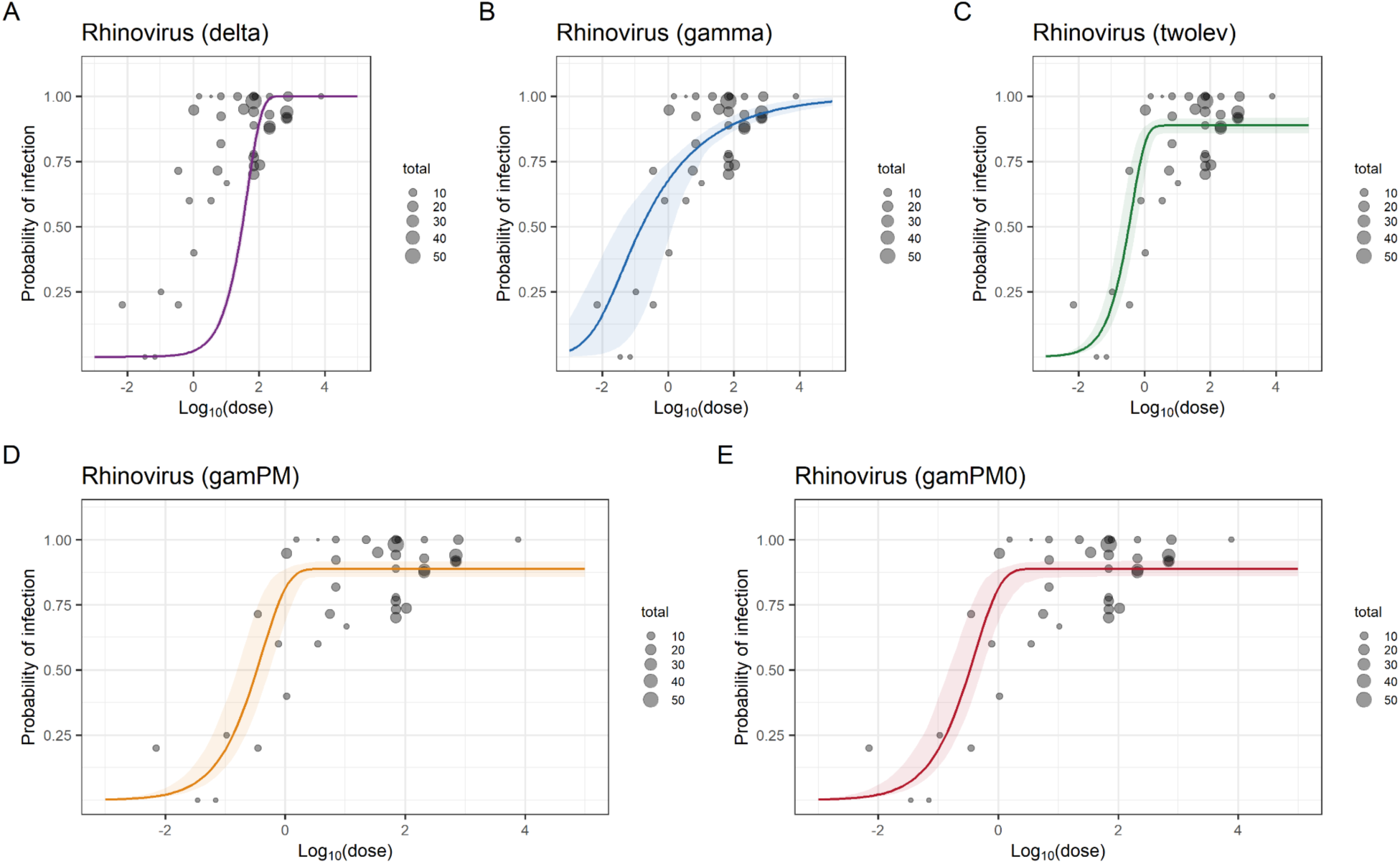
Fitted dose response curves for rhinovirus based on observed human challenge data. Grey bubbles represent observed data, with circle size proportional to the number of participants in each trial at each dose level. Colored curves show fitted dose–response models. Panels display each model for rhinovirus: (A) dirac delta model; (B) gamma model; (C) two-level model; (D) gamma plus point-mass model; (E) gamma plus point-mass at zero model.

**Figure S7.**
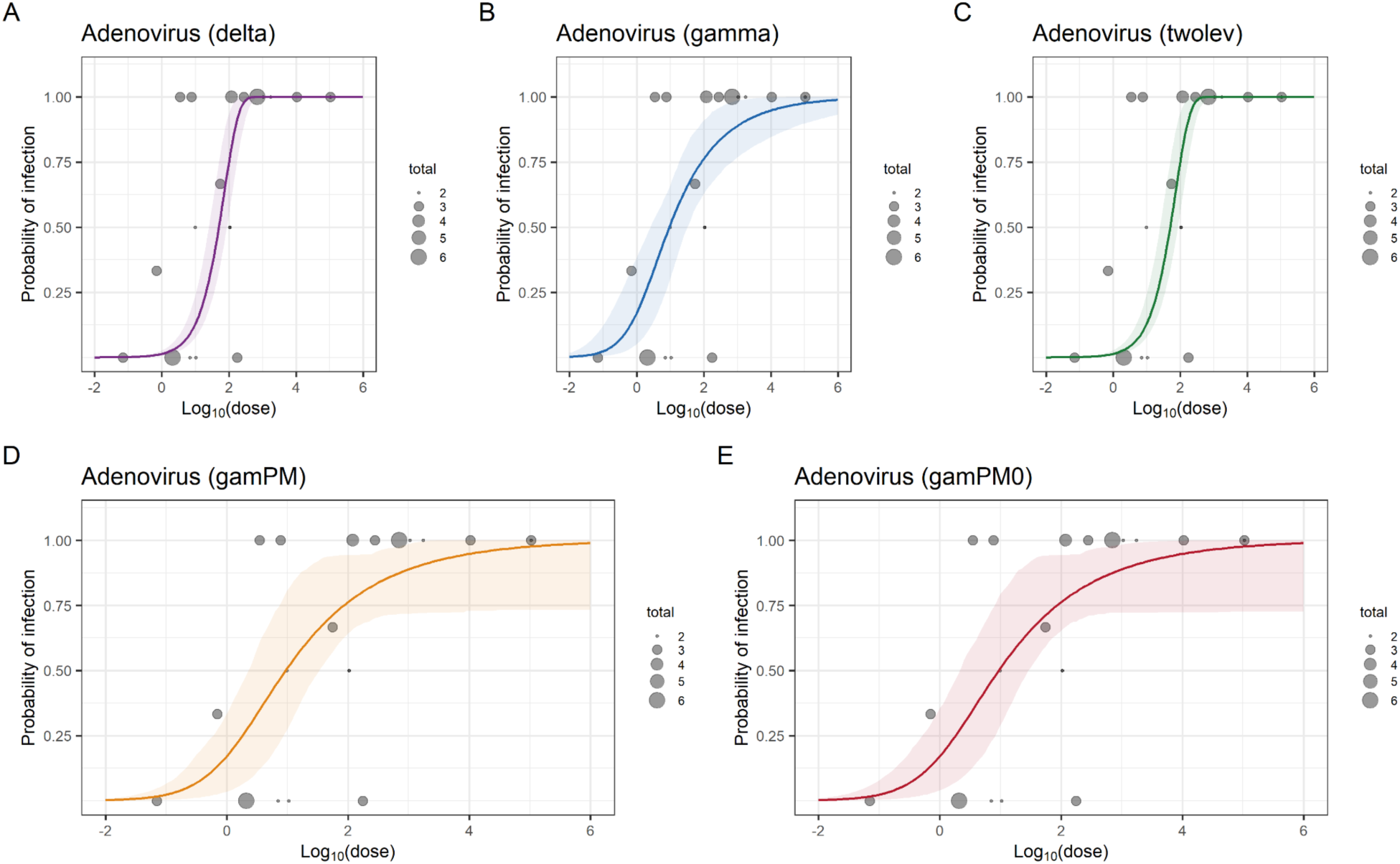
Fitted dose response curves for adenovirus based on observed human challenge data. Grey bubbles represent observed data, with circle size proportional to the number of participants in each trial at each dose level. Colored curves show fitted dose–response models. Panels display each model for adenovirus: (A) dirac delta model; (B) gamma model; (C) two-level model; (D) gamma plus point-mass model; (E) gamma plus point-mass at zero model.

**Figure S8.**
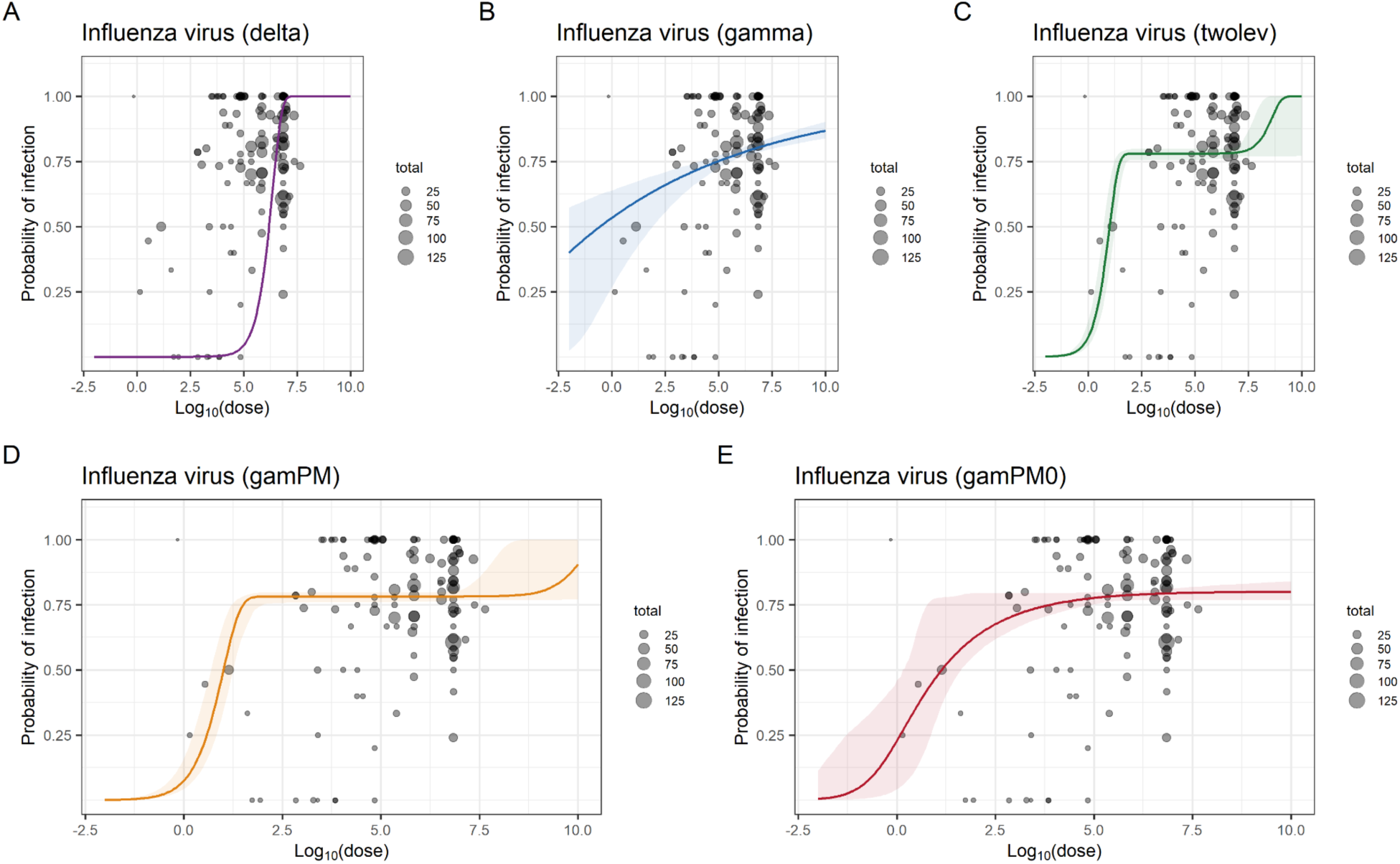
Fitted dose response curves for influenza virus based on observed human challenge data. Grey bubbles represent observed data, with circle size proportional to the number of participants in each trial at each dose level. Colored curves show fitted dose–response models. Panels display each model for influenza virus: (A) dirac delta model; (B) gamma model; (C) two-level model; (D) gamma plus point-mass model; (E) gamma plus point-mass at zero model.

**Figure S9.**
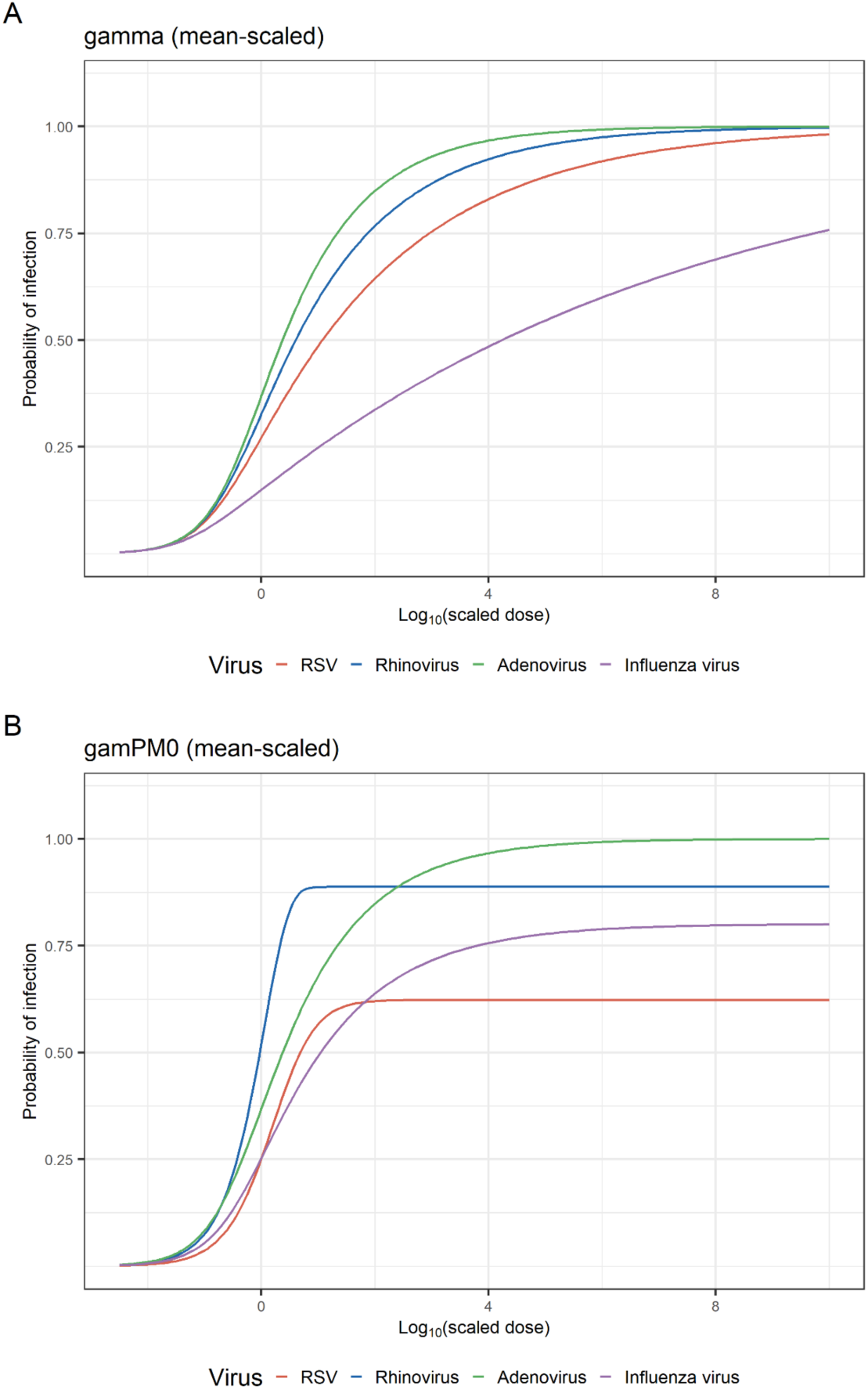
Fitted mean-scaled dose response curves for each pathogen based on observed human challenge data. Colored curves show each pathogen (red: RSV; blue: rhinovirus; green: adenovirus; purple: influenza virus). Panels display the fitted dose response models: (A) mean-scaled gamma plus point-mass model; (B) mean-scaled gamma plus point-mass at zero model.

## Reference

1. Baker RE, Park SW, Yang W, Vecchi GA, Metcalf CJE, Grenfell BT. The impact of COVID-19 nonpharmaceutical interventions on the future dynamics of endemic infections. Proc Natl Acad Sci U S A. 2020 Dec;117(48):30547–53. doi:10.1073/pnas.2013182117 PubMed PMID: 33168723; PubMed Central PMCID: PMC7720203.

2. Bents SJ, Martin ET, Stevens-Ayers T, Andrews C, Adler A, Perofsky AC, et al. Multiplex serology reveals age-specific immunodynamics of respiratory pathogens in the wake of the COVID-19 pandemic. Nat Commun. 2025 Dec 10;16(1):11015. doi:10.1038/s41467-025-65957-9

3. Jamrozik E, Selgelid MJ. History of Human Challenge Studies. Human Challenge Studies in Endemic Settings. 2020 Aug 19;9–23. doi:10.1007/978-3-030-41480-1_2 PubMed PMID: null; PubMed Central PMCID: PMC7431914.

4. Oxford JS, Catchpole A, Mann A, Bell A, Noulin N, Gill D, et al. A Brief History of Human Challenge Studies (1900-2021) Emphasising the Virology, Regulatory and Ethical Requirements, Raison D’etre, Ethnography, Selection of Volunteers and Unit Design. Curr Top Microbiol Immunol. 2024;445:1–32. doi:10.1007/82_2022_253 PubMed PMID: 35704095.

5. Adams-Phipps J, Toomey D, Więcek W, Schmit V, Wilkinson J, Scholl K, et al. A Systematic Review of Human Challenge Trials, Designs, and Safety. Clin Infect Dis. 2023 Feb;76(4):609–19. doi:10.1093/cid/ciac820 PubMed PMID: 36219704; PubMed Central PMCID: PMC9938741.

6. Teunis PF, Nagelkerke NJ, Haas CN. Dose response models for infectious gastroenteritis. Risk Anal. 1999 Dec;19(6):1251–60. doi:10.1023/a:1007055316559 PubMed PMID: 10765461.

7. Gomes MGM, Lipsitch M, Wargo AR, Kurath G, Rebelo C, Medley GF, et al. A missing dimension in measures of vaccination impacts. PLoS Pathog. 2014 Mar;10(3):e1003849. doi:10.1371/journal.ppat.1003849 PubMed PMID: 24603721; PubMed Central PMCID: PMC3946326.

8. Nilsen V, Wyller J. QMRA for Drinking Water: 1. Revisiting the Mathematical Structure of Single-Hit Dose-Response Models. Risk Analysis. 2016;36(1):145–62. doi:10.1111/risa.12389

9. Miura F, Klinkenberg D, Wallinga J. Quantifying the Individual Variation in Susceptibility to Endemic Coronavirus and SARS-CoV-2 with Human Challenge Trials. Epidemiology. 2024 Jan;35(1):113–7. doi:10.1097/EDE.0000000000001679 PubMed PMID: 38032803; PubMed Central PMCID: PMC10683973.

10. Haas CN, Rose JB, Gerba CP. Quantitative Microbial Risk Assessment [Internet]. Wiley; 2014. Available from: https://onlinelibrary.wiley.com/doi/book/10.1002/9781118910030 doi:10.1002/9781118910030

11. Miura F, Klinkenberg D, Ainslie KEC, Backer JA, Leung KY, McDonald SA, et al. Inferring vaccine efficacy and mode of action from human challenge studies [Internet]. medRxiv; 2023 [cited 2026 May 28]. p. 2023.09.24.23296054. Available from: https://www.medrxiv.org/content/10.1101/2023.09.24.23296054v1 doi:10.1101/2023.09.24.23296054

12. Teunis PF, Havelaar AH. The Beta Poisson dose-response model is not a single-hit model. Risk Anal. 2000 Aug;20(4):513–20. doi:10.1111/0272-4332.204048 PubMed PMID: 11051074.

13. Messner MJ, Berger P, Nappier SP. Fractional poisson--a simple dose-response model for human norovirus. Risk Anal. 2014 Oct;34(10):1820–9. doi:10.1111/risa.12207 PubMed PMID: 24724739.

14. Lambkin-Williams R, Noulin N, Mann A, Catchpole A, Gilbert AS. The human viral challenge model: accelerating the evaluation of respiratory antivirals, vaccines and novel diagnostics. Respir Res. 2018;19:123. doi:10.1186/s12931-018-0784-1 PubMed PMID: 29929556; PubMed Central PMCID: PMC6013893.

15. Carter J, Saunders V. Virology: principles and applications. 1st Edition. Wiley; 2007.

16. Kim JH, Liepkalns J, Reber AJ, Lu X, Music N, Jacob J, et al. Prior infection with influenza virus but not vaccination leaves a long-term immunological imprint that intensifies the protective efficacy of antigenically drifted vaccine strains. Vaccine. 2016 Jan 20;34(4):495–502. doi:10.1016/j.vaccine.2015.11.077 PubMed PMID: 26706277; PubMed Central PMCID: PMC4713344.

17. Gould VMW, Francis JN, Anderson KJ, Georges B, Cope AV, Tregoning JS. Nasal IgA Provides Protection against Human Influenza Challenge in Volunteers with Low Serum Influenza Antibody Titre. Front Microbiol. 2017 May 17;8:900. doi:10.3389/fmicb.2017.00900 PubMed PMID: 28567036; PubMed Central PMCID: PMC5434144.

18. Chaumont A, Martin A, Flamaing J, Wiseman DJ, Vandermeulen C, Jongert E, et al. Host immune response to respiratory syncytial virus infection and its contribution to protection and susceptibility in adults: a systematic literature review. Expert Rev Clin Immunol. 2025 Jun;21(6):745–60. doi:10.1080/1744666X.2025.2494658 PubMed PMID: 40278893.

19. Stobart CC, Nosek JM, Moore ML. Rhinovirus Biology, Antigenic Diversity, and Advancements in the Design of a Human Rhinovirus Vaccine. Front Microbiol. 2017;8:2412. doi:10.3389/fmicb.2017.02412 PubMed PMID: 29259600; PubMed Central PMCID: PMC5723287.

20. Abo YN, Jamrozik E, McCarthy JS, Roestenberg M, Steer AC, Osowicki J. Strategic and scientific contributions of human challenge trials for vaccine development: facts versus fantasy. Lancet Infect Dis. 2023 Dec;23(12):e533–46. doi:10.1016/S1473-3099(23)00294-3 PubMed PMID: 37573871.

